# Xylazine Spreads Across the US: A Growing Component of the Increasingly Synthetic and Polysubstance Overdose Crisis

**DOI:** 10.1101/2021.09.20.21263680

**Authors:** Joseph Friedman, Fernando Montero Castillo, Phillippe Bourgois, Rafik Wahbi, Daniel Dye, David Goodman, Chelsea Shover

## Abstract

**Background:** Recent sharp exacerbations of the US overdose crisis have been linked to systemic polysubstance use and potent synthetic compounds in numerous drug classes. Xylazine is a veterinary tranquilizer, long noted in the opioid supply of Puerto Rico, and more recently Philadelphia. Yet its national growth over time, geographic distribution, and potential role in the shifting US overdose risk environment are poorly characterized.

**Methods:** In this sequential mixed methods study, xylazine was increasingly observed by our ethnographic team over many years of intensive participant observation fieldwork in Philadelphia among drug sellers and people who inject drugs (PWID). Subsequently, we systematically searched for records describing xylazine-involved overdose mortality across the US and assessed time trends and overlap with other drugs.

**Results:** In 10 jurisdictions—representing all 4 US Census Regions—xylazine was found to be increasingly implicated in overdose mortality, rising from 0.36% of deaths in 2015 to 6.7% in 2020. The highest xylazine prevalence in the recent data was observed in Philadelphia, (25.8% of deaths), followed by Maryland (19.3%) and Connecticut (10.2%). Illicitly-manufactured-fentanyls were present in 98.4% of xylazine-involved-overdose-deaths— suggesting a strong ecological link—as well as cocaine (45.4%), benzodiazepines (28.4%), heroin (23.3%), and alcohol (19.7%). PWID in Philadelphia described xylazine as a sought-after adulterant that improves euphoria and lengthens the short duration of fentanyl injections, in particular. They also linked it to increased risk of soft tissue infection and overdose.

**Conclusions:** We summarize evidence that xylazine is increasingly implicated in overdose deaths across the US and is linked to the proliferation of illicitly-manufactured-fentanyls. We document hypothesis-generating ethnographic accounts linking it to health risks for PWID. Nevertheless, many jurisdictions do not routinely test for xylazine, and it is not tracked in federal overdose statistics. Further efforts are needed to provide PWID with services that can help to minimize additional risks associated with a shifting drug supply.

## INTRODUCTION

The US overdose crisis has accelerated exponentially for the past four decades, with a shifting profile of substances driving fatalies^1^. From 2000-2006 cocaine was the leading drug associated with overdose deaths, which was replaced successively by prescription opioids (2007-2013), heroin (2014-2015) and illicitly-manufactured fentanyls (2016-present)^1^. In recent years, sharp increases in fatalities have been linked to systemic polysubstance use and potent synthetic compounds in numerous drug classes, including synthetic opioids such as fentanyls, sedatives, stimulants such as methamphetamine, and novel benzodiazepines^2–6^. As overdose rates continue to reach unprecedented heights^7^, there is a need to continue to examine novel synthetic compounds, and polysubstance use patterns, which are increasingly implicated.

Xylazine is a veterinary tranquilizer, which is not approved for human use in the United States, but is commonly used for sedating large animals^4,8^. Although human intoxication with xylazine has been reported sporadically over the past several decades in a number of case studies^8,9^, it was first described as a more prevalent additive in the unregulated drug supply of Puerto Rico^4,10,11^. It was also noted in the literature describing drug-overdose deaths in Philadelphia as early as 2006, yet it did not appear in high prevalence at that time^12^. However, since the mid-2010s xylazine has been noticed by people who inject drugs (PWID) and public health practitioners as an increasingly commonplace additive in the street opioid supply of Philadelphia^13^. Further, recent reports from Connecticut implicated xylazine in a rising fraction of overdose deaths in 2019-2020^14,15^. A report released in September 2021 leveraged data from 38 states and Washington DC representing the year 2019, and found xylazine to be present in 1.8% of overdose deaths^16^. However, no time trends were provided, and results were not disaggregated below the level of US Census Region, with limits the usefulness of the results for frontline providers and harm reductionists. Additionally, reports from Philadelphia and Connecticut, as well as media reports from numerous cities, suggest that increases in xylazine-involved overdose have increased sharply in 2020-2021. Therefore, additional study with more recent and detailed geographic information is urgently needed to assess the national relevance and trajectory of xylazine-involved overdose fatalities, and their role in the rapidly shifting US overdose risk environment. Additionally, further qualitative information based on the perspectives of people who inject drugs (PWID) is needed to better understand why xylazine might be spreading across the US, and the potential health and other risks involved.

In the current sequential mixed methods analysis, over many years of participant observation fieldwork in Philadelphia (2007-2021) among drug sellers and people who inject drugs (PWID) in the Puerto Rican inner city, xylazine was frequently referred to as a powerful adulterant/enhancer of heroin, “back in Puerto Rico.” In the mid to late 2010s, suddenly, Xylazine appeared in the Philadelphia fentanyl-dominated opioid supply, rendering it “difficult to obtain real heroin” according to street-based PWID. Subsequently, we systematically searched for records describing xylazine-involved overdose mortality across the US and assessed time trends and overlap with other drugs.

## METHODS

### Ethnographic methods

Beginning in 2007 our initial ethnographic team (PB and FM, later JF) set out to study urban poverty, violence, and drug markets in majority Puerto Rican neighborhoods in North Philadelphia^17–19^. Leveraging a longitudinal ethnographic approach^20,21^ to studying these topics, we rented an apartment adjacent to several cocaine and heroin sales points, in the heart of Philadelphia’s sprawling open-air narcotics economy. Various members of the ethnographic team lived full-time in our field site for different periods, and all have since made periodic follow-up visits to conduct extensive interviews with longitudinal informants through the present. This has allowed us to participate in the local social scene as neighbors, and study drug markets by triangulating multiple local perspectives (drug sellers, drug consumers, residents, harm reduction providers, law enforcement) impacted by drug markets. This long-term triangulated immersion reduces social desirability bias, allowing for the comparison of self-reported practices beliefs and observation of real-time practices of stigmatized, illegal hard-to-document activities such as injection drug use, drug sales, and violence. With IRB approval, and full informant consent, we tape-recorded hundreds of interviews, and wrote detailed observational fieldnotes. Most interviews were conducted informally in a conversational format, often in street-settings, while accompanying respondents in their normal neighborhood activities. Over many years we developed genuine friendships with many of our respondents, which we still enjoy. This facilitates long-term follow-up with difficult to reach, stigmatized, and structurally vulnerable populations. In particular, ethnographic immersion in drug use subcultures allows for real-time documentation of shifts in illicit drug markets and their multifactored potential implications for health risks, which often cannot be ascertained from epidemiological records alone.

Our ethnographic database from Philadelphia consists of nearly 1,500 pages of fieldnotes as well as transcriptions from hundreds of interviews, dozens of hours of video, and thousands of photographs. We loaded all textual data into the NVivo qualitative analysis platform and coded for emergent themes and characters. Analyses were conducted iteratively, throughout the fieldwork process, with emerging analytical insights further refining follow-up questions, and targeting follow-up research activities (see ^17,18,20–22^). Qualitative data gathered during 15 years of ethnographic fieldwork generated numerous quantitative hypotheses, which we have explored through multimethod collaborations with quantitative researchers^17,23,24^.

### Quantitative methods

Quantitative data describing overdose deaths were collected as part of a larger research effort^25^ to assemble and assess granular person-level records from jurisdictions that publish overdose data ahead of federal statistics (which are typically available in their finalized form on a 12-24 month lag^26^). Although xylazine was not described in existing national-level records^26^ at the time of writing this article, its presence in overdose deaths could be ascertained from the cause of death fields in records obtained from medical examiner and coroner offices from numerous jurisdictions. Therefore, we assessed our existing database of data from 10 jurisdictions for the presence of xylazine implicated in overdose deaths. Further, we systematically searched for additional jurisdictions where xylazine positivity has been noted, which we identified through popular press reports and official statistical reports. Wherever possible, we requested and incorporated person-level records describing xylazine deaths from each jurisdiction, as well as the overlap with other drugs. In some cases, only aggregated reports were available.

We summarized xylazine-involved overdose deaths per jurisdiction and year, both as counts, and as a rate per reported overdose deaths. For a small number of jurisdictions, the denominator (all overdose deaths) was not available from the same source as the numerator (xylazine-involved deaths) and was obtained from federal statistics^26,27^. As this analysis was completed in September 2021, in some cases data for part of 2021 were available. In order to provide standardized metrics, counts of xylazine-involved deaths and total overdose deaths for 2021 were estimated by assuming that both would continue linearly for the remainder of 2021. For jurisdictions where drug overlaps could be assessed, we visualized clusters of drugs found to co-occur with xylazine using UpSet visualization^28^. All quantitative analyses were conducted using R version 4.0.3.

## RESULTS

### Quantitative results

In overdose data from 10 jurisdictions—representing all 4 major US census regions—Xylazine was found to be increasingly implicated in overdose mortality (Figure 1). The highest prevalence was observed in Philadelphia, (with xylazine implicated in 25.8% of overdose deaths in 2020), followed by Maryland (19.3% in 2021) and Connecticut (10.2% in 2020). In 2021, xylazine prevalence also grew substantially in Jefferson County, Alabama, reaching 8.4% of overdose fatalities. Across the 4 census regions, the Northeast had the highest prevalence, and the West had the lowest (Figure 2), with only 6 xylazine-involved overdose deaths in total detected in Phoenix, Arizona, and 1 in San Diego County, California. Across jurisdictions, a clear increasing trend was noted. Pooling data for 2015, a total xylazine prevalence of 0.36% was observed. By 2020, this had grown to 6.7% of overdose deaths, representing a 20-fold increase. From 2019 to 2020—the last year of data available for all jurisdictions—the prevalence increased by 44.8%.

**Figure 1.**
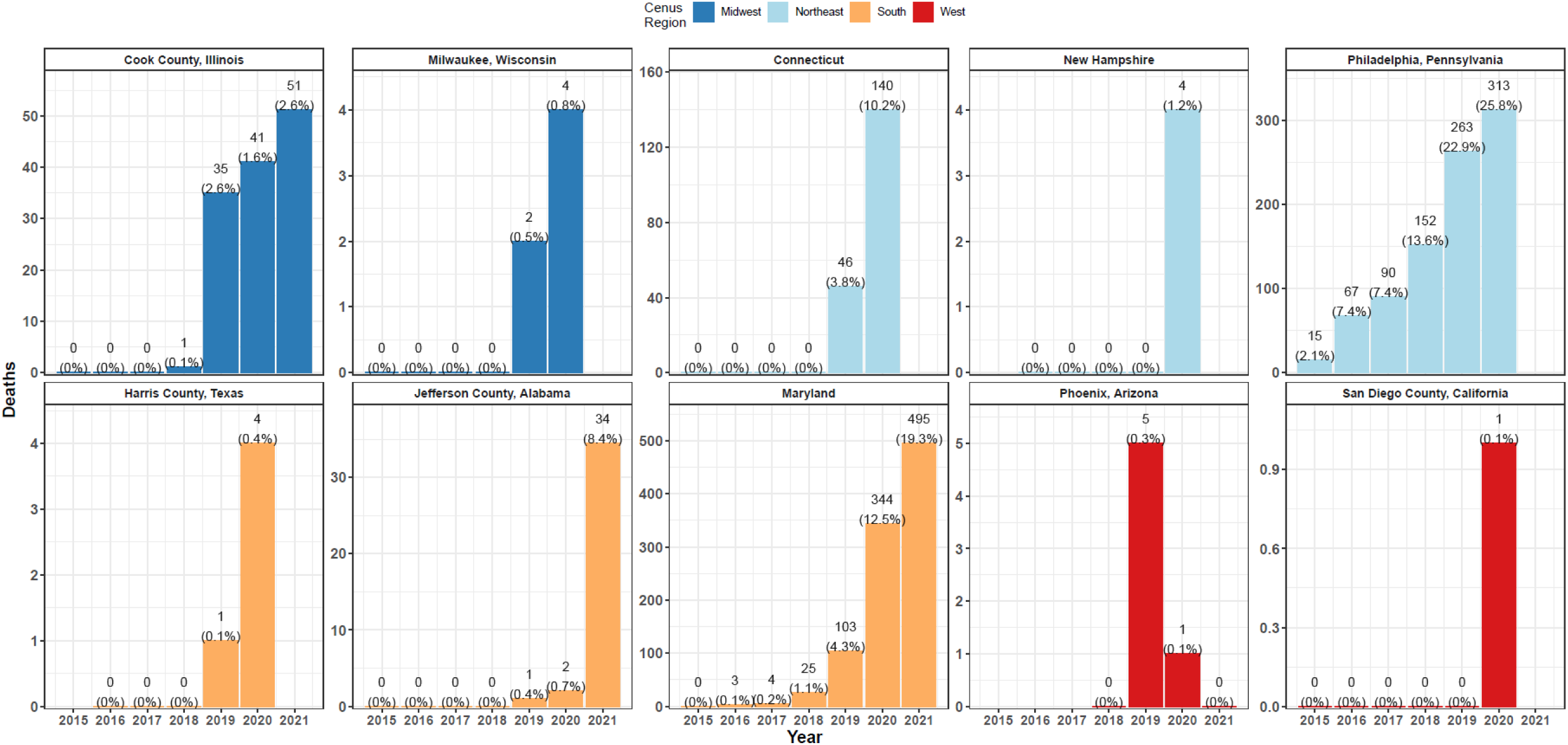
Xylazine-Involved Overdose Deaths by Jurisdiction and Year. Xylazine-involved deaths are show as counts and as a percent of all overdose deaths in text. Color indicates US census region. Values for 2021 represent estimates, should trends from the observed fraction of the year continue linearly.

**Figure 2.**
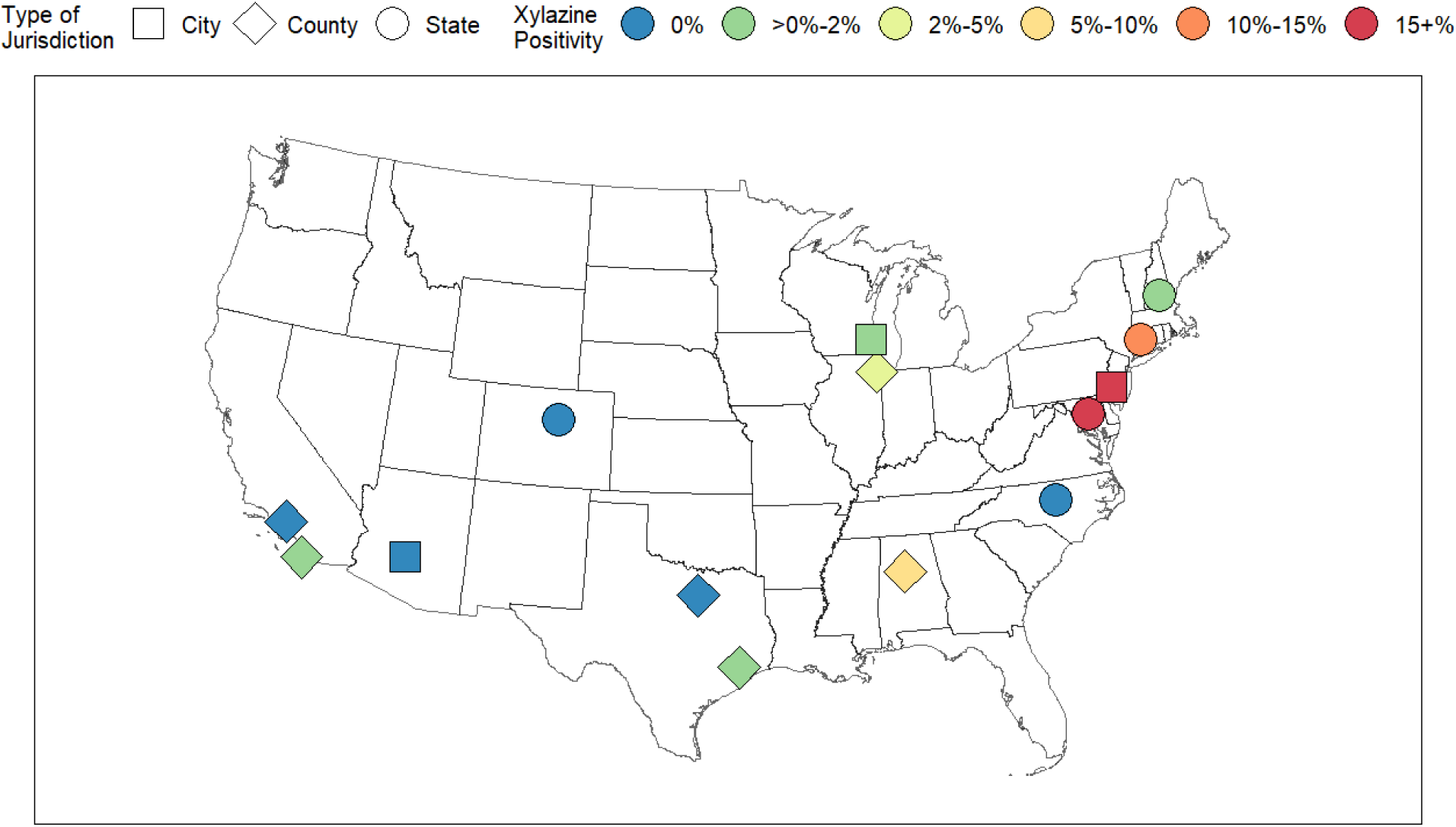
Geographic Distribution of Xylazine Positivity in Overdose Deaths. This figures summarizes the geographic distribution of xylazine positivity in overdose deaths in the full database of 14 locations. Point shape corresponds to type of jurisdiction. Color corresponds to the magnitude of xylazine positivity in the most recent year of data available for each location. Values for 2021 represent estimates, should trends from the observed fraction of the year continue linearly. The time period shown in each point can be seen in Supplemental Table 1.

In four jurisdictions, xylazine was not found to be implicated in overdose deaths, including: Phoenix, Arizona; Tarrant County, Texas; Denver, Colorado; and North Carolina (Figure 2, and Supplemental Table 1). However, in North Carolina, xylazine has been detected in drug checking data^29^, and state officials confirmed that it has been found in toxicological testing during death investigations. However, it is not yet reflected in death certificates related to these cases.

Among the jurisdictions with a substantial number of xylazine deaths, and where full drug overlaps could be assessed (including Connecticut; Cook County, Illinois; Jefferson County, Alabama; and Philadelphia, Pennsylvania) xylazine was observed to co-occur with all 6 drug classes assessed (Figure 3). Fentanyl was the most commonly co-occurring drug, involved in 98.4% of xylazine-involved overdoses, suggesting a strong ecological relationship. Cocaine (45.3%), benzodiazepines (28.4%), heroin (23.3%), and alcohol (19.7%) were also common. Methamphetamine was the least frequently observed overlapping drug, implicated in only 10.0% of xylazine-involved overdose deaths.

**Figure 3.**
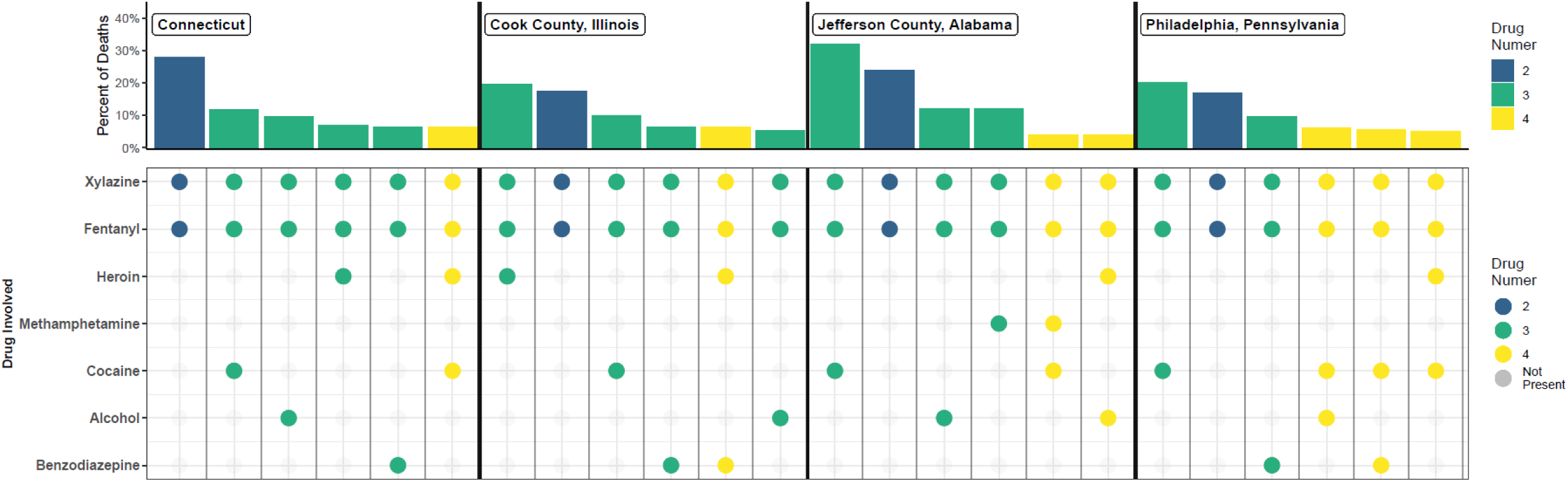
Drug Overlap in Xylazine-Involved Overdose Deaths. Data are shown for the jurisdictions where drug overlaps with xylazine could be determined, for the 6 most commonly observed drug combinations in each jurisdiction. Each column corresponds to one cluster of drug classes. The drugs present in each cluster of deaths is indicated in the bottom panel, by the presence of a solid dot. The top panel shows the percent of all xylazine-involved overdose deaths in that jurisdiction pertaining to each cluster. Color indicates the total number of drug classes implicated in each cluster.

Substantial variation was found between jurisdictions in the most common drug combinations observed (Figure 3). In Jefferson County, Alabama, and Philadelphia, Pennsylvania, xylazine was most commonly found with both fentanyl and cocaine, representing 32.0% and 20.0%, respectively, of all overdose deaths in those jurisdictions. In Cook County, Illinois, xylazine was most commonly found with fentanyl, and heroin, representing 19.6% of cases. In Connecticut, xylazine and fentanyl were most commonly found alone, constituting 28.0% of cases.

### Ethnographic results

#### Popular conception of xylazine prior to introduction - Connection to Puerto Rico

Prior to the widespread availability of xylazine in the Philadelphia drug supply, it was often mentioned in passing by residents of the majority Puerto Rican neighborhood where our fieldwork was based as a powerfully psychoactive additive ‘“back on the Island”.’ Xylazine was occasionally detected in fatal overdoses in Philadelphia as early as 2006^12^, but it was not common knowledge among PWID. Significantly, however, many of our long-term informants recently immigrating/returning from Puerto Rico spoke with a mix of intrigue and apprehension about the psychoactive effects and health risks of “*anastesia de caballo* [horse tranquilizer]”. As early as 2009 xylazine was already beginning to achieve notoriety among PWID in Puerto Rican Philadelphia:

> *“Pero la droga aquí es muy diferente a la de Puerto Rico…[but the drugs here are very different from what they’re using in Puerto Rico] over there they’re lacin’ it with that stuff…horse tranquilizer, what they call anestesia de caballo…for like 8 years now…and they inject it, and you see them fast asleep on the street corner. And also, they say that that when they shoot up and skin it, you know your skin gets, like, kind of fucked up and you get these big craters. And those craters over there, I mean, they’re not like the ones that the dopefiends from here got*.*”*

#### Introduction of xylazine to Philadelphia drug scene

At least a decade after Xylazine became a fixture in Puerto Rico, it entered the street opioid supply in Philadelphia as a more prevalent additive in the mid-2010s. The shift was noted by PWID, as well as harm reductionists and city public health officials^13^. PWID began to describe xylazine—often referred to as *tranq*—as a known element of specific ‘stamps’ or brands of opioid products in the illicit retail market. Opioid formulations containing xylazine, (e.g.,”*tranq dope”*) became largely sought-after, as the addition of xylazine was reported to improve the euphoria and prolong the duration of fentanyl injections, in particular, solving “the problem” of the “short legs” of the otherwise euphoric effects of illicitly manufactured fentanyl. Nevertheless, a ratio of too much xylazine to fentanyl was said to distort the initial injection “rush” and euphoric “nod” from opioids, and leave a consumer overly sedated:

> *“FM: Have you noticed changes in the heroin since you started using?*
>
> *Definitely. Here it’s all tranqy, it’s all tranq*.
>
> *FM: Do you like the tranq dope?*

*Yeah I like a good tranq-fent bag. Some people don’t like it, but plenty of people do. It’s sought after. Certain stamps [brands printed on drug bags] that are known for tranq have better business, you know what I mean? I like it cuz, fentanyl is such a short-lived high… It’s a good high, but it’s so short that the nod is over real quick and you get sicker faster. See, the tranq like extends the high, it gives the dope more of a heroin effect, it’s a good rush with the heroin-like effect. But then other times, they straight put bags out there that are just all tranq. You shoot it, you feel no rush, and you’re just out, you’re asleep for at least 40 minutes. You’re sitting there one second talking, and then you’re waking up 2-3 hours later in a weird position*.*”*

Despite often expressing positive sentiments for the psychoactive potency of opioid-xylazine combinations, PWID frequently expressed concern about novel health risks from ‘*tranq dope’*. In particular, concerns related to increased risk of soft tissue damage:

> *“I got some friends out here that got really torn up by it, you know they got holes in them, abscesses, basically it’s like the body is rotting. People here are losing limbs like with gangrene. Whatever they’re doing with the tranq…*.*everybody is getting these scabby sores all over their bodies… and many of them don’t shoot meth. So it’s from the dope. You know what I mean, you had the dope, then the fentanyl, now it’s the “tranq-fent,” the rhinoceros tranquilizer, the horse tranquilizer, you know? Cooked up and it’s broken down and it’s added to the dope and we seek that out, you know what I mean? Our habits are fentanyl and tranquilizer*.*”*

For other PWID, concerns about the negative health effects of xylazine outweighed interest in its psychoactive potency. Further, some individuals expressed a strong preference for “real heroin” or fentanyl-heroin formulations, not containing xylazine, based on a different character of the euphoric effect:

> *“There’s all kinds of weird cuts that are coming up now. You really can’t know what you’re getting these days. I stay with the same people every time I buy, to try to limit some of that. So I go over to [name redacted] because it’s pretty good and it never has any tranq in it. Tranq causes people to black out, and causes amnesia, people do dangerous stuff they wouldn’t normally do like walk in front of a car, and they don’t even realize what’s going on, they’re in a whole other world. And if you fuck it up and miss [the vein], your whole arm turns black. Oddly enough, I would think that nobody would want to use it, but some people specifically seek bags that have tranq in it, or what they think is tranq (laughing)*.
>
> *FM: Have you used it?*
>
> *Yeah accidentally. It’s dangerous stuff. I was doing like zombie walks from the tranq, so I was walking, but I wasn’t aware of what was going on, and I fell on the train tracks and cracked my skull. I literally cracked my skull open, and almost died. So yeah. You know, the old heroin, that was just heroin, was dangerous enough, but the tranq is just really a whole other world*.*”*

Harm reductionists and frontline medical providers also noted increased frequency and severity of injection-related soft tissue damage. One harm reductionist noted:

> *“People had been talking about it [xylazine] as a cutting agent or as a way of cheating drug users for a long time. But the intentional addition of high amounts of xylazine to help give fentanyl legs [increase the duration of effect] really started increasing since maybe 2019. And that’s when we started to see way more people coming in with necrotizing skin and soft tissue issues. The amount of medical complaints related to xylazine was pretty astounding and terrifying. Xylazine wounds are a whole other kind of…just horror. And they were really exacerbated by the shift to fentanyl, since folks were injecting more and more frequently, so with each one of the injections, if there’s xylazine in it, and if you miss [the vein], you were risking a really deep and necrotizing wound*.*”*

Xylazine was also linked by frontline providers to potentially increased, or shifting profiles of overdose risk:

> *“JF: what’s your sense of overdose risk with xylazine?*
>
> *The number of people I was reversing [administering naloxone] was definitely going up. There were people who would become only minimally alert and be only slightly responsive to Narcan. So that definitely concerned us that something different was happening, that there were non-opioid agents involved. We did a lot of Narcan training, and we had to focus more on the necessity of doing rescue breaths, instead of just Narcan*.*”*

PWID also noted that xylazine presence in street drug formulations could be distinguished by a characteristic taste that can be observed immediately after injection:

> “*You know a bag got tranq in it because you’ll shoot it and your mouth goes dry right away, and you know, you taste it*.*”*

#### Xylazine in an evolving drug supply

It is important to note that the widespread introduction of xylazine into the drug supply of Philadelphia occurred at a time of numerous other shifts, which may complicate efforts to pinpoint the effects of xylazine on population health^30^. Of particular note, methamphetamine grew in popularity, prevalence, and ease of access in the years leading up to 2021:

> *“FM: And is it easy to find meth around here?*
>
> *Yeah. When we first started coming, back around like 2 years ago, it wasn’t anywhere, nobody even knew what it was really. And now it’s everywhere, it’s just everywhere. I’d say on like ¾ of the corners they’re selling it. They’re starting to give [free] samples of it now*.*”*

At the same time, the market in Philadelphia was in a longer-term process of shifting to an increasingly illicitly-manufactured-fentanyl-based opioid supply. PWID celebrated a highly visible ramification of this shift: the steep drop in the unit price of ‘heroin’. Whereas a packet of heroin had a retail street price of $10 USD for the first decade of our fieldwork, the price suddenly dropped to $5 USD in the late 2010s, as fentanyl began to dominate the market^30^. At the same time, this also prompted PWID to increase the number of bags purchased each day, especially given the much shorter half-life of fentanyl analogues compared to traditional heroin^2^. This led to a sharp increase in the number of injections per day for the average consumer of street opioids.

The unregulated drug supply at the end of the 2010s and early 2020s in Philadelphia was undergoing a process of extensive experimentation with prepackaged polysubstance combinations. Ratios of heroin to illicitly-manufactured fentanyl were experimented with, as well as the addition of other psychoactive substances such as other benzodiazepines, xylazine, and other additives. At times customers would be unaware of these additions, yet in other moments the alleged composition of polysubstance mixtures served as a point of street-based marketing to boost sales. In response, PWID became skilled at navigating a constantly shifting drug supply to maximize the potency per dollar spent:

> *FM: Do you go to the same corner every time, or do you switch it up?*
>
> *Well, we usually ask around like “what’s good”. Because even if they give samples in the morning and it’s good stuff, when you go back to buy it it’s not the same stuff. So you’ve gotta ask everyone who’s doing it around you, “What’s good? What’s good?” and then whatever they say is good right now, that’s where we go. There’s a couple spots where you can still get real heroin. Any $10 bag is usually gonna be straight heroin, straight dope. At [street name redacted], they’re mixed, it’s real dope but there’s some fentanyl in it too. An’ they cut it with Xanax, they put some benzos in. Everybody’s got like, their own little flair, what they do with their shit*.

Similarly, harm reductionists emphasized that xylazine became commonplace in the context of a rapidly shifting drug supply, especially with respect to the replacement of heroin and prescription opioids with illicitly-manufactured fentanyls:

> *I would say that Xylazine is another trend in concert with K2 [synthetic cannabinoids] in Philly. Part of the reason that people use so much K2 here is because there’s this belief that cannabinoids in general increase the half-life of opioids. When the switch to fentanyl happened, drug users were talking about how frustrating it is that fentanyl doesn’t have legs. Manufacturers responded by trying to increase the amount of sedation, by adding xylazine. And likewise, people think that K2 can help ‘give fentanyl legs’ [increase the duration of effect]. So xylazine is really part of a larger story of both drug users and dealers adapting to the new world of fentanyl in both safe and unsafe ways*.

## DISCUSSION

We summarize longitudinal, recent, and geographically specific evidence describing how xylazine is increasingly implicated in overdose deaths in jurisdictions spanning all major US regions and link it to detailed ethnographic observations of its use in Philadelphia open-air narcotics markets. Xylazine presence in overdose deaths grew exponentially during the observed period, rising nearly 20-fold between 2015 and 2020. Whereas the most recent national data from the State Unintentional Drug Overdose Reporting System characterized the level of xylazine-involved overdoses in 2019^16^, we found that the prevalence increased by nearly 50% from 2019 to 2020 alone, indicating a need for more recent data to guide the public health response. Nevertheless, we find that even looking at only 10 jurisdictions a greater number of xylazine-involved overdose deaths were seen in 2020 (854), than the previous study looking at 38 states in 2019 (826)^16^, implying a very fast rate of growth nationally.

Xylazine prevalence was observed earliest and at the highest magnitude in the Northeast, and may be spreading west, in a pattern similar to the trajectory of illicitly-manufactured fentanyls in recent years^25^. This similarity may not be incidental, as an analysis of the co-occurrence of fentanyl and xylazine indicates a strong ecological link, with fentanyl nearly universally implicated in xylazine-involved overdose deaths. Further, ethnographic data among PWID suggests that the use of xylazine as an illicit drug additive may predominantly serve as a response to the short duration of fentanyl. By ‘giving fentanyl legs’—offering improved duration of effect—the addition of xylazine may confer a competitive market advantage for illicit opioid formulations that contain it, as it remedies one of the most commonly expressed complaints that PWID hold regarding fentanyl-based street opioid formulations.

Ethnographic data offer a potential explanation for Philadelphia’s early adoption of xylazine in the drug supply, relative to other regions of the US. We observed that, for at least a decade before Xylazine became a widely used additive to the street opioid supply in Puerto Rican-dominated Philadelphia open-air drug markets, there was an existing commonplace conceptualization of it as a potent, exciting, and dangerous drug, based on local return migration ties to Puerto Rico. This may help explain why Philadelphia appears to be the earliest, and largest-magnitude documented emerging epicenter of xylazine use in the United States. Additionally, it is noteworthy that the retail narcotics economy in Philadelphia is dramatically concentrated in the city’s Puerto Rican neighborhoods^17^. For example, based on geocoded data from the Philadelphia police department, the lowest-income Puerto Rican neighborhoods in Philadelphia have a concentration of ‘narcotics violations’ over ten-fold higher than the most impoverished majority non-Hispanic Black neighborhoods of the city^17^. Similarly, in ethnographic fieldwork it was clear that majority Puerto Rican neighborhoods are host to remarkably higher-volume and more profitable retail narcotics markets compared to any other part of the city^17,18^. This has been hypothesized to reflect the unique function of majority-Puerto Rican neighborhoods as a more phenotypically diverse meeting ground in an otherwise heavily residentially segregated city, where majority Black and White areas are typically clearly demarcated^17^. Puerto Rican cultural norms continue to play numerous key roles in Philadelphia’s retail drug market, and Puerto Rican drug sellers control a large fraction of the street opioid sale^18,19^. In sum, the Philadelphia street opioid scene has numerous strong ties to Puerto Rico that may have influenced the early entrance of xylazine in Philadelphia, given an existing understanding and precedent for its use (See ^17–19,30^).

Ethnographic data also link the entrance of xylazine into the drug supply to novel health risks for PWID. In particular, both PWID and harm reductionists in Philadelphia reported a new prevalence and magnitude of soft tissue damage, which has also been described in the literature from Puerto Rico, where xylazine has had a longer-term presence^4,8^. Various mechanisms have been hypothesized linking xylazine use to increased soft tissue injury, including necrosis stemming from localized tissue hypoxemia, and reduce sensitivity to skin injury^4,10^. Xylazine was also linked in ethnographic accounts to the potential for increased overdose risk. As xylazine is known to cause hypotension and bradycardia, these effects may be synergistic with opioid agonists, exacerbating overdose risk^14^. Further, naloxone may not fully reverse overdose symptoms from opioid-xylazine formulations, given that naloxone does not act on non-opioid sedatives^13,14^. Nevertheless, we note that xylazine gained popularity in Philadelphia—and the United States more generally—during a particularly complicated and multifactorial moment for the shifting drug supply. For example, the entrance of xylazine co-occurred with the increased polysubstance use of methamphetamine, numerous synthetic opioids, novel benzodiazepines, and synthetic cannabinoids, which could all exert overlapping influences on health risks. Careful study will be needed to parse out the health implications of each of these co-occurring factors.

The spread of xylazine across the US illustrates the increasingly synthetic and polysubstance-use-oriented nature of the US overdose crisis, which has profound implications for epidemiologic surveillance of overdose mortality. Improved efforts are needed to ensure that novel substances identified in overdose deaths anywhere in the US can be quickly added to toxicological testing across the approximately 2,000 medical examiner and coroner jurisdictions in the US^31^. As many jurisdictions are currently not routinely testing for xylazine in overdose fatalities, the magnitude of its epidemiological significance may be misunderstood at present. In the current data-sparse landscape about polysubstance drug formulations and overdose patterns, new efforts to provide routine, nuanced testing results from samples of drugs and syringes offer great value for guiding continuous improvements of surveillance sytems^32–34^. Ideally, nuanced drug checking results assessing for the presence of various novel substances could be quickly linked to widespread toxicological analysis for death investigation at the national level. This is currently quite challenging given the fractured nature of the US medical examiner and coroner system. However, the increased usage of centralized laboratories, either publicly or privately, represents a promising avenue. Further, federal data collection and reporting platforms^26^ must be modified to convey the nuanced combinations of drugs driving the modern iteration of the US overdose crisis. Currently reported deaths can only be viewed separately by drug class, with impedes the assessment of polysubstance overdose deaths nationally. Streamlining the detection of novel substances, and integration into toxicological testing through reporting systems represents a key challenge, but an essential one for the rapid detection of overdose risks.

The limitations of this study are numerous and especially related to the limited nature of public, recent data pertaining to xylazine in the US drug supply. We provide data from only a sample of jurisdictions, which do not represent the wider US population. No publicly available data are available from the majority of the country, and we are therefore likely to be vastly underestimating the scope of xylazine entrance to the drug supply in terms of absolute numbers. As a rate of all overdose deaths, our non-representative could be biased up or down relative to the underlying—and unknown—national xylazine prevalence. Additionally, our ethnographic data and analysis should be regarded as mainly hypothesis generating, as they reflect data from a specific subset of PWID and harm reductionists from one city. It is also important to note that little data is available to track the non-fatal implications of xylazine, especially soft tissue damage, which was a major area of concern for PWID informants.

In sum, we found evidence of a national signal that xylazine is prevalent in the unregulated drug supply and increasing in specific jurisdictions. This growth is likely to have epidemiological significance in the coming years similar to the spread of fentanyl. This has profound implications for the evolving risk environment for PWID in the US. Renewed efforts are needed to improve surveillance for novel substances and sharing of this data to researchers, policymakers, harm reductionists and people who use substances. Granular drug testing results with a more rapid turnaround are needed to avert the growing overdose epidemic. More urgently, PWID must be provided with resources that can help to minimize additional risks associated with a shifting and increasingly synthetic and polysubstance drug supply with drug formulation-specific harm reduction advice and services.

## Data Availability

The quantitative data used in this study are publicly available upon request from each of the medical examiner or coroner jurisdictions represented. However, the authors are not permitted to distribute the records. The authors may be contacted for assistance in requesting records from data providers.

---

**Supplemental Table 1.** Data Availability. This figures summarizes data availability across the 14 jurisdictions assessed. Values for 2021 represent estimates, should trends from the observed fraction of the year continue linearly. *Although xylazine is absent in official records from North Carolina, it has been detected in toxicological testing and drug checking data from the state.

| Jurisdiction | Type | Region | Most Recent Year | Xylazine Deaths | Percent of Overdose Deaths |
| --- | --- | --- | --- | --- | --- |
| Philadelphia, Pennsylvania | City | Northeast | 2020 | 313 | 25.8% |
| Maryland | State | South | 2021 | 495 | 19.3% |
| Connecticut | State | Northeast | 2020 | 140 | 10.2% |
| Jefferson County, Alabama | County | South | 2021 | 34 | 8.4% |
| Cook County, Illinois | County | Midwest | 2021 | 51 | 2.6% |
| New Hampshire | State | Northeast | 2020 | 4 | 1.2% |
| Milwaukee, Wisconsin | City | Midwest | 2020 | 4 | 0.8% |
| Harris County, Texas | County | South | 2020 | 4 | 0.4% |
| San Diego County, California | County | West | 2020 | 1 | 0.1% |
| Phoenix, Arizona | City | West | 2021 | 0 | 0.0% |
| Tarrant County, TX | County | South | 2021 | 0 | 0.0% |
| Denver, Colorado | State | West | 2019 | 0 | 0.0% |
| Los Angeles County, CA | County | West | 2020 | 0 | 0.0% |
| North Carolina | State | South | 2020 | 0* | 0.0%* |

